# A prospective study of risk factors associated with seroprevalence of SARS-CoV-2 antibodies in healthcare workers at a large UK teaching hospital

**DOI:** 10.1101/2020.11.03.20220699

**Authors:** Daniel J Cooper, Sara Lear, Laura Watson, Ashley Shaw, Mark Ferris, Rainer Doffinger, Rachel Bousfield, Katherine Sharrocks, Michael P. Weekes, Ben Warne, Dominic Sparkes, Nick K Jones, Lucy Rivett, Matthew Routledge, Afzal Chaudhry, Katherine Dempsey, Montgomery Matson, Adil Lakha, George Gathercole, Olivia O’Connor, Emily Wilson, Orthi Shahzad, Kieran Toms, Rachel Thompson, Ian Halsall, David Halsall, Sally Houghton, Sofia Papadia, Nathalie Kingston, Kathleen E Stirrups, Barbara Graves, Neil Walker, Hannah Stark, the CITIID-NIHR BioResource COVID-19 Collaboration, Daniela De Angelis, Shaun Seaman, John R Bradley, M. Estée Török, Ian Goodfellow, Stephen Baker

**Affiliations:** Cambridge University Hospitals NHS Foundation Trust, Cambridge, UK; Global and Tropical Health Division, Menzies School of Heath Research and Charles Darwin University, Darwin, Northern Territory, Australia; NIHR Cambridge Clinical Research Facility; Clinical Microbiology and Public Health Laboratory, Public Health England, United Kingdom; University of Cambridge School of Clinical Medicine, Cambridge, UK; NIHR BioResource, NIHR Cambridge Biomedical Research Centre, Cambridge Biomedical Campus, Cambridge, UK; Department of Public Health and Primary Care, School of Clinical Medicine, University of Cambridge, Cambridge Biomedical Campus, Cambridge, UK; Department of Haematology, School of Medicine, University of Cambridge, Cambridge Biomedical Campus, Cambridge, UK; MRC Biostatistics Unit, University of Cambridge, Cambridge, UK; Department of Medicine, University of Cambridge School of Clinical Medicine, Cambridge Biomedical Campus, Cambridge, UK; Department of pathology, Division of virology, University of Cambridge, Cambridge Biomedical Campus, Cambridge, UK; Cambridge Institute of Therapeutic Immunology and Infectious Disease, University of Cambridge School of Clinical Medicine, Cambridge Biomedical Campus, Cambridge, UK

**Author notes:** corresponding author: Dr Daniel J Cooper. Senior authors (Ian Goodfellow, Stephen Baker).

**Keywords:** SARS-CoV-2, COVID-19, sero-epidemiology, healthcare workers, risk factor analysis

## Abstract

**Background:** The COVID-19 pandemic continues to grow at an unprecedented rate. Healthcare workers (HCWs) are at higher risk of SARS-CoV-2 infection than the general population but risk factors for HCW infection are not well described.

**Methods:** We conducted a prospective sero-epidemiological study of HCWs at a UK teaching hospital using a SARS-CoV-2 immunoassay. Risk factors for seropositivity were analysed using multivariate logistic regression.

**Findings:** 410/5,698 (7·2%) staff tested positive for SARS-CoV-2 antibodies. Seroprevalence was higher in those working in designated COVID-19 areas compared with other areas (9·47% versus 6·16%) Healthcare assistants (aOR 2·06 [95%CI 1·14-3·71]; *p*=0·016) and domestic and portering staff (aOR 3·45 [95% CI 1·07-11·42]; *p*=0·039) had significantly higher seroprevalence than other staff groups after adjusting for age, sex, ethnicity and COVID-19 working location. Staff working in acute medicine and medical sub-specialities were also at higher risk (aOR 2·07 [95% CI 1·31-3·25]; *p*<0·002). Staff from Black, Asian and minority ethnic (BAME) backgrounds had an aOR of 1·65 (95% CI 1·32 – 2·07; *p*<0·001) compared to white staff; this increased risk was independent of COVID-19 area working. The only symptoms significantly associated with seropositivity in a multivariable model were loss of sense of taste or smell, fever and myalgia; 31% of staff testing positive reported no prior symptoms.

**Interpretation:** Risk of SARS-CoV-2 infection amongst HCWs is heterogeneous and influenced by COVID-19 working location, role, age and ethnicity. Increased risk amongst BAME staff cannot be accounted for solely by occupational factors.

**Funding:** Wellcome Trust, Addenbrookes Charitable Trust, National Institute for Health Research, Academy of Medical Sciences, the Health Foundation and the NIHR Cambridge Biomedical Research Centre.

**Research in context:** 

**Evidence before this study:** Specific risk factors for SARS-CoV-2 infection in healthcare workers (HCWs) are not well defined. Additionally, it is not clear how population level risk factors influence occupational risk in defined demographic groups. Only by identifying these factors can we mitigate and reduce the risk of occupational SARS-CoV-2 infection. We performed a review of the evidence for HCW-specific risk factors for SARS-CoV-2 infection. We searched PubMed with the terms “SARS-CoV-2” OR “COVID-19” AND “Healthcare worker” OR “Healthcare Personnel” AND “Risk factor” to identify any studies published in any language between December 2019 and September 2020. The search identified 266 studies and included a meta-analysis and two observational studies assessing HCW cohort seroprevalence data. Seroprevalence and risk factors for HCW infections varied between studies, with contradictory findings. In the two serological studies, one identified a significant increased risk of seroprevalence in those working with COVID-19 patients (Eyre *et al* 2020), as well as associations with job role and department. The other study (Dimcheff *et al* 2020) found no significant association between seropositivity and any identified demographic or occupational factor. A meta-analysis of HCW (Gomez-Ochoa *et al* 2020) assessed >230,000 participants as a pooled analysis, including diagnoses by both RT-PCR and seropositivity for SARS-CoV-2 antibodies and found great heterogeneity in study design and reported contradictory findings. Of note, they report a seropositivity rate of 7% across all studies reporting SARS-CoV-2 antibodies in HCWs. Nurses were the most frequently affected healthcare personnel and staff working in non-emergency inpatient settings were the most frequently affected group. Our search found no prospective studies systematically evaluating HCW specific risk factors based entirely on seroprevalence data.

**Added value of this study:** Our prospective cohort study of almost 6,000 HCWs at a large UK teaching hospital strengthens previous findings from UK-based cohorts in identifying an increased risk of SARS-CoV-2 exposure amongst HCWs. Specifically, factors associated with SARS-CoV-2 exposure include caring for confirmed COVID-19 cases and identifying as being within specific ethnic groups (BAME staff). We further delineated the risk amongst BAME staff and demonstrate that occupational factors alone do not account for all of the increased risk amongst this group. We demonstrate for the first time that healthcare assistants represent a key at-risk occupational group, and challenge previous findings of significantly higher risk amongst nursing staff. Seroprevalence in staff not working in areas with confirmed COVID-19 patients was only marginally higher than that of the general population within the same geographical region. This observation could suggest the increased risk amongst HCWs arises through occupational exposure to confirmed cases and could account for the overall higher seroprevalence in HCWs, rather than purely the presence of staff in healthcare facilities. Over 30% of seropositive staff had not reported symptoms consistent with COVID-19, and in those who did report symptoms, differentiating COVID-19 from other causes based on symptom data alone was unreliable.

**Implications of all the available evidence:** International efforts to reduce the risk of SARS-CoV-2 infection amongst HCWs need to be prioritised. The risk of SARS-CoV-2 infection amongst HCWs is heterogenous but also follows demonstrable patterns. Potential mechanisms to reduce the risk for staff working in areas with confirmed COVID-19 patients include improved training in hand hygiene and personal protective equipment (PPE), better access to high quality PPE, and frequent asymptomatic testing. Wider asymptomatic testing in healthcare facilities has the potential to reduce spread of SARS-CoV-2 within hospitals, thereby reducing patient and staff risk and limiting spread between hospitals and into the wider community. The increased risk of COVID-19 amongst BAME staff cannot be explained by purely occupational factors; however, the increased risk amongst minority ethnic groups identified here was stark and necessitates further evaluation.

## Background

With >30 million cases and >1 million deaths reported to date globally, the ongoing COVID-19 pandemic continues to impact daily life^1^. A nationwide lockdown in the UK on 23^rd^ March 2020 succeeded in slowing infection rates^2^; however, a rising number of infections are now being observed with the easing of lockdown measures^3^. The logistics of managing patients with COVID-19 presented unique challenges to hospitals and NHS trusts across the UK; evidence and practices evolved rapidly as experience was gained. Healthcare workers (HCWs) are at a higher risk of SARS-CoV-2 infection than the general population^4,5^, and some contradictory evidence is emerging for risk factors associated with SARS-CoV-2 infection in front-line HCWs^6-8^. Protecting HCWs by identifying risk factors for SARS-CoV-2 infection as the UK continues to see an increase in COVID-19 diagnoses is paramount^3^. Controlling transmission within a hospital setting, as well as from hospitals back into the community, is key for controlling the pandemic^9,10^. However, defining HCW specific risk-factors remains a challenge. Additionally, higher rates of symptomatic SARS-CoV-2 infection, hospitalisation and death have been observed amongst patients from ethnic minority populations in the UK^11^ and worldwide^12,13^; the reasons for this disparity are unclear. Reported infections in HCW suggests higher mortality in HCWs from minority backgrounds^14^; however, it is not yet clear to what extent workplace exposures influence infection. Here, we present the results of a large sero-epidemiological study of SARS-CoV-2 seropositivity in staff at a teaching hospital in the East of England.

## Methods

### Setting

Cambridge University Hospitals NHS Foundation Trust (CUH) is a tertiary referral centre and teaching hospital with 1,000 beds and 11,545 staff serving a population of 580,000 people in the East of England. The facility is equipped with a 20-bed ICU, a 23-bed neurosciences and trauma ICU, and an Emergency Department that receives ∼14,000 attendees a month. Between March and June 2020, CUH treated 525 patients with PCR-confirmed COVID-19 (**Figure 1**). The peak of COVID-19 admissions occurred in late March and early April 2020, with comparatively few COVID-19 admissions from June 2020 onwards. The definition of COVID-19 working for the purpose of risk stratification included clinical areas designated as either “Red” (patients with PCR-confirmed SARS-CoV-2 infection) or “Amber” (patients for whom there is a high clinical suspicion of COVID-19).

**Figure 1:**
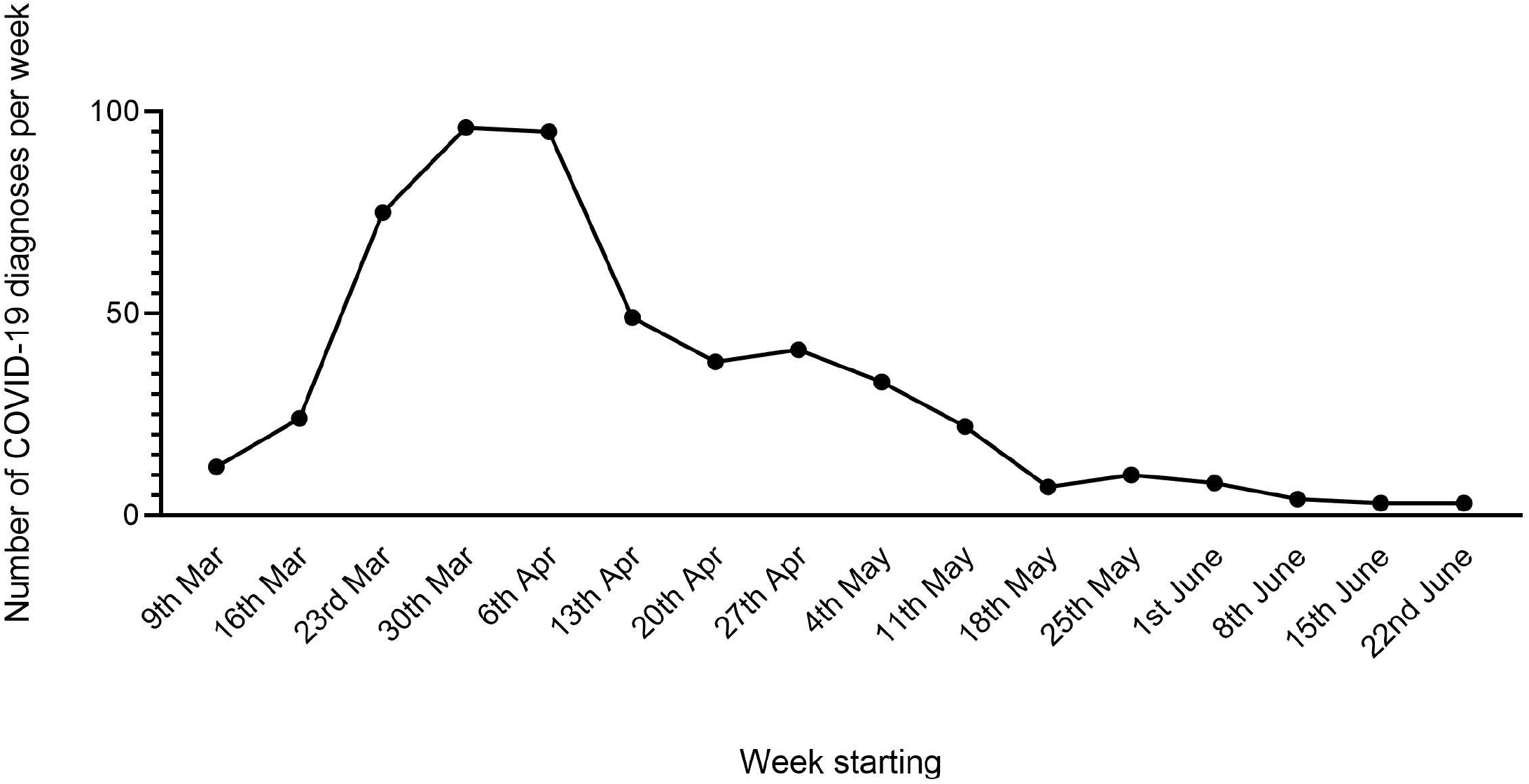
Epidemic curve of COVID-19 admissions at Cambridge University Hospitals NHS Foundation Trust.

As of September 2020, the East of England reported 27,516 laboratory-confirmed cases of SARSCoV-2 infection^15^, with a corresponding population rate of 441·2 per 100,000 people to date. This rate is substantially less than the worst affected regions of North West England (772·9/100,000) and Yorkshire and the Humber (693·2/100,000)^15^. According to the 2011 England and Wales census^16^ 85·3% of the population of the East of England are White British, 5·5% are White Other, 4·8% are Asian, 2% are Black, and 1·9% are of Mixed ethnicity. The proportion of Black, Asian, and Minority Ethnic (BAME) staff employed at CUH is representative of the overall NHS workforce^17^ (21·2% vs 20·7%, respectively).

An asymptomatic staff screening programme using SARS-CoV-2 PCR testing was established in April 2020^18^. A staff screening programme for SARS-CoV-2 serological testing was commenced on the 10^th^ June 2020. All staff members were invited by email to participate in the serological screening programme and asked to self-refer for a clinic appointment. Written informed consent was obtained from all participants. As part of this process all participants were invited to join the NIHR BioResource – COVID-19 Research Cohort (IRAS 220277). At enrolment, participants completed a questionnaire asking about demographic characteristics, healthcare role, ethnicity, previous symptoms consistent with COVID-19 and previous results of SARS-CoV-2 PCR testing. A total of 7·8 ml of blood was collected, including one serum sample and one whole blood sample. The serum sample was assayed for total SARS-CoV-2 antibody; both residual serum and whole blood were stored for future analyses.

### Laboratory assays

Serological testing for antibodies directed against SARS-CoV-2 was performed using the Centaur XP SARS-Cov-2 Total Antibody assay (Siemens Healthcare Limited, Surrey, UK). This method is a fully automated high throughput enzyme linked chemiluminescent bridging immunoassay which targets the S1RBD antigen of SARS-CoV-2 and can detect all Ig subclasses (IgG, IgM, and IgA). The quantity of SARS-CoV2 antibodies correlates directly with relative light units (RLU), which is converted to Index Values with a measuring interval of 0.05 ->10 index, where values below 1 are reported as nonreactive and those ≥1.0 are reported as reactive, as validated by the manufacturer by clinical correlation. The method was independently validated by Public Health England and has a reported sensitivity and specificity of 98.1% (95% CI 96.6 – 99.1) and 99.9% (95% CI 99.4 – 100)^19^ respectively. Samples were processed in the Biochemistry laboratory at CUH following the SOP as stated by the manufacturer in their Instruction for Use (IFU) after a local verification using guidance from The Royal College of Pathologists^20^.

As previously described, the RT-PCR assay used at CUH designates a cycle threshold (Ct) of ≤36 to correspond to a positive result^18^.

### Statistical analysis

Seroprevalence is reported as a percentage ([proportion with antibodies/number tested] x 100). Logistic regression was used for univariable and multivariable analyses of seroprevalence comparisons. The Wilcoxon rank-sum test was used for comparison of median Ct values. Data were analysed using Stata v14.2 (StataCorp, College Station, Texas).

### Ethical approval

Ethical approval for this study was granted by the East of England – Cambridge Central Research Ethics Committee (IRAS ID: 220277).

## Findings

### Baseline information

The CUH staff serology screening clinic was operational between 10^th^ June and 7 August 2020. A total of 8,376 (73%) staff attended the clinic for SARS-CoV-2 serology; 5,697/8,376 (68%) of these consented to be enrolled in the study (Figure 2). 1,700/5,967 (28·5%) of study participants reported that they had worked in a designated COVID-19 area within the CUH structure during the peak of the epidemic between February and June 2020. The median age of participants was 38 years (range 17-83 years) and 22·7% (1,293/5,697) were male (Table 1). A total of 22 staff required hospital admission for COVID-19. No CUH staff members died.

**Table 1:**
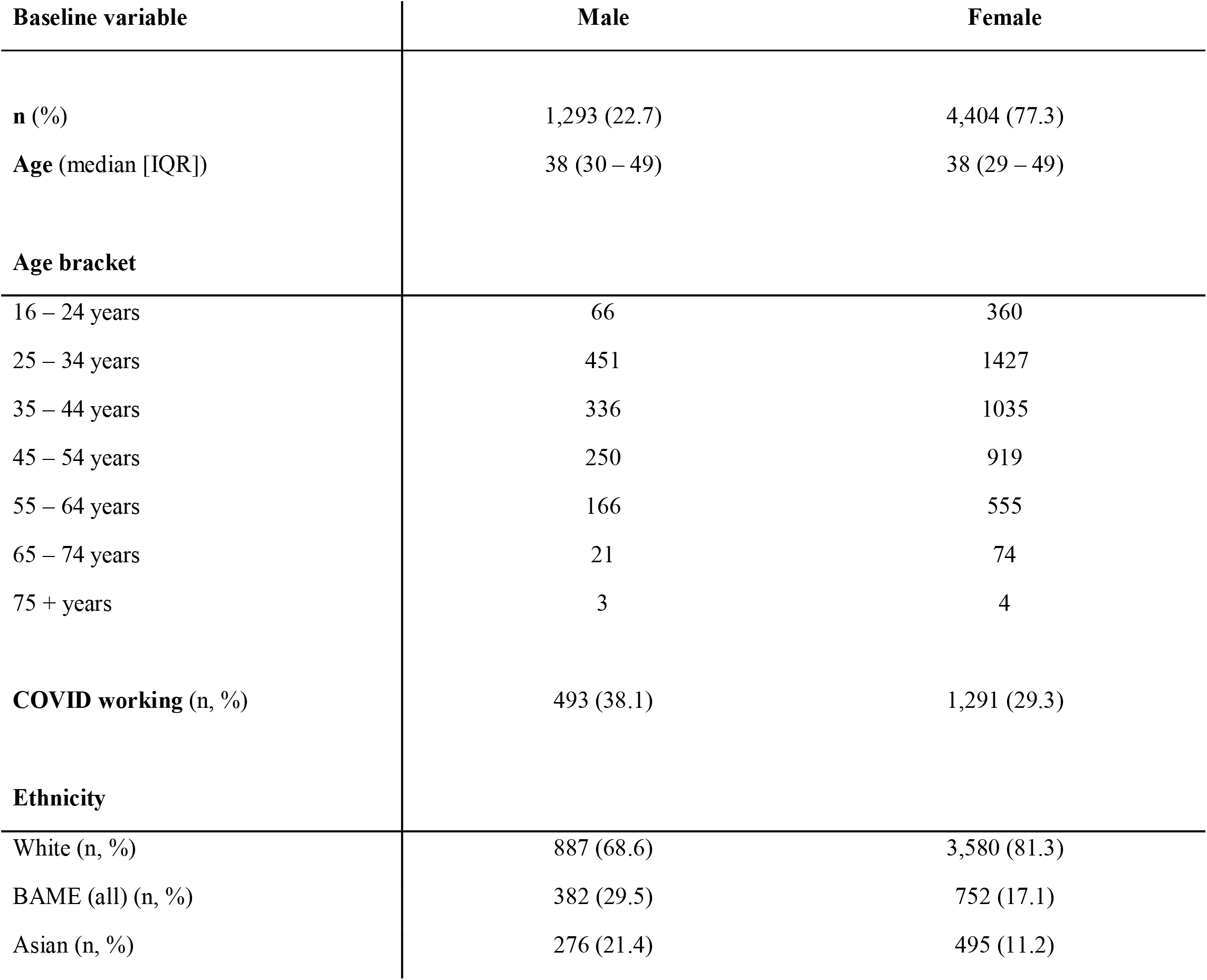
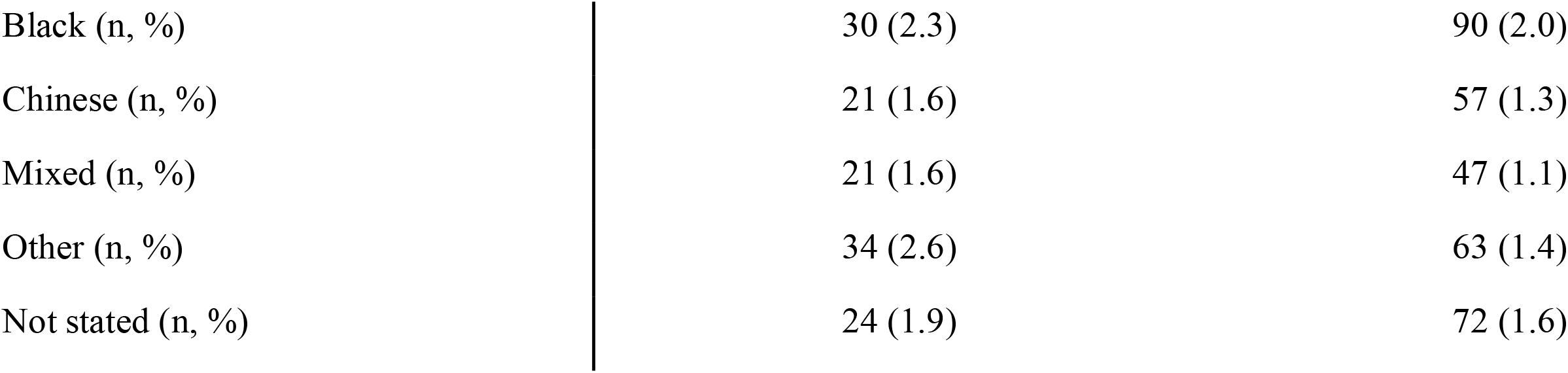
Baseline demographics

**Figure 2:**
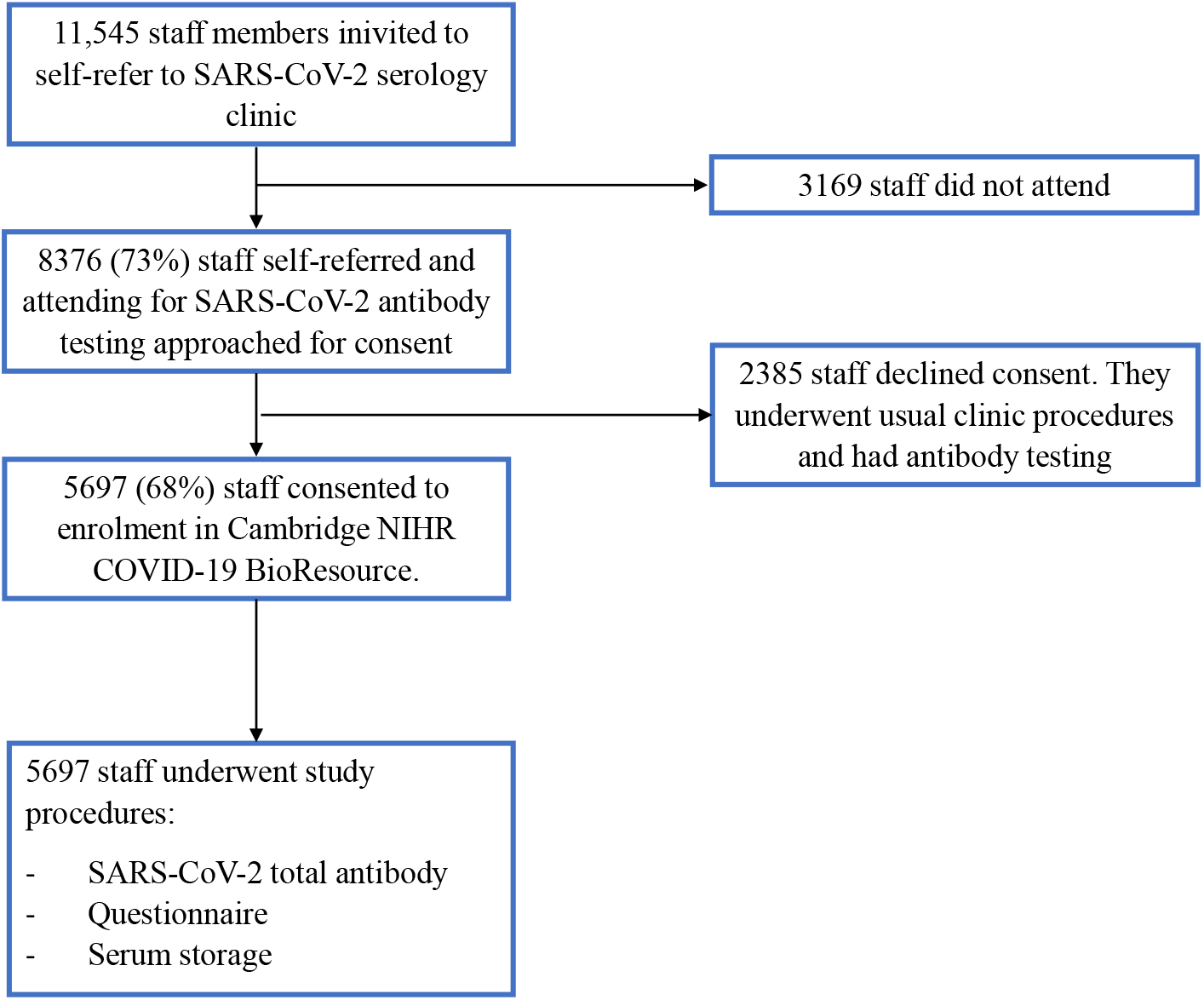
Study flowchart.

### Seroprevalence

The overall seroprevalence of total SARS-CoV-2 antibodies amongst all staff in this study was 7·2% (410/5,698). Amongst those reporting having worked in a dedicated COVID-19 area between February and June 2020, the seroprevalence increased to 9·5% (169/1,784; Table 2). Conversely, the comparable seroprevalence in those reporting they had never worked in COVID-19 area was 6·2% (241/3,913; *p*<0·0001). The prevalence of seropositivity in male staff (8·0%; 104/1293) was not significantly different (*p*=0·18) than that observed in female staff (6·95%; 306/4404). The risk of seropositivity decreased with age, with an odds ratio (OR) of 0·83 (95% CI 0·76 – 0·91) for every 10-year increase in age (*p*<0·0001).

**Table 2:**
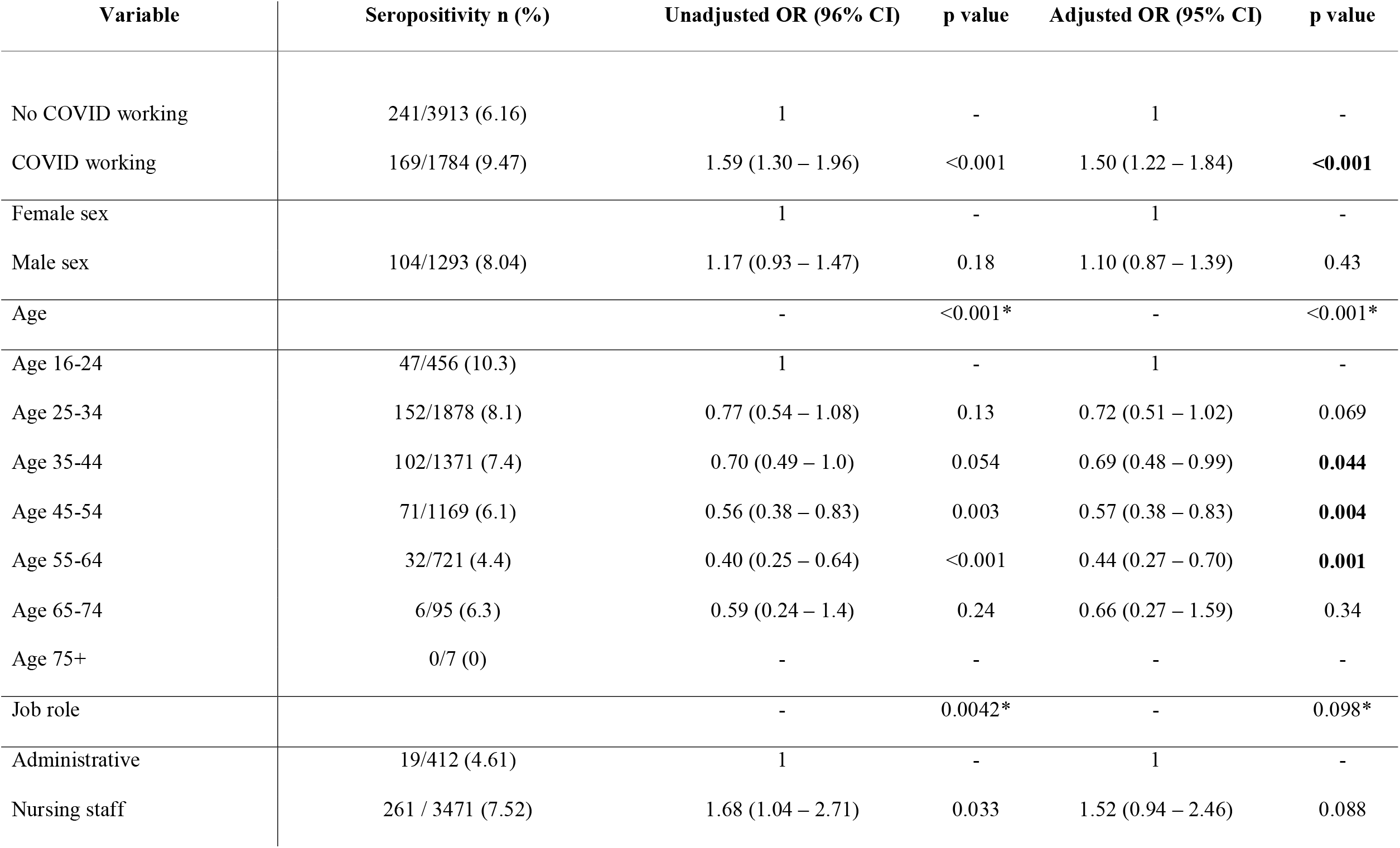
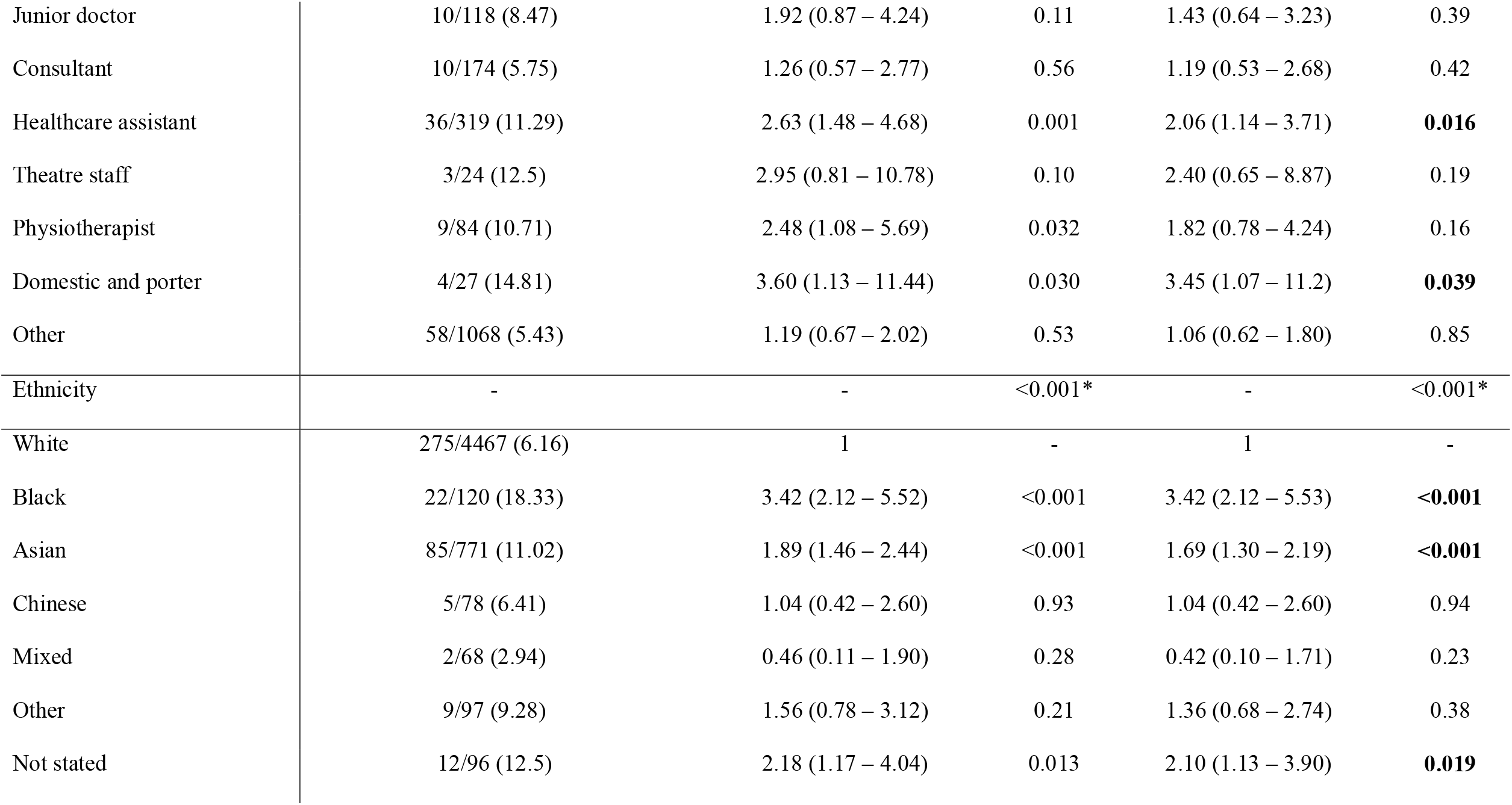
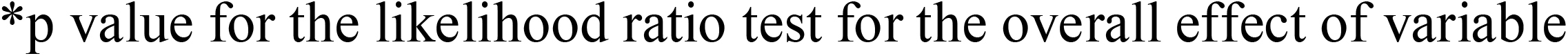
Odds ratio (OR) and adjusted odds ratio (aOR) for variables associated with seropositivity aORs calculated using a multivariable model containing serostatus, age, sex, ethnicity, job role and COVID-working location.

| Variable | Seropositivity n (%) | Unadjusted OR (96% CI) | p value | Adjusted OR (95% CI) | p value |
| --- | --- | --- | --- | --- | --- |
| No COVID working | 241/3913 (6.16) | 1 | - | 1 | - |
| COVID working | 169/1784 (9.47) | 1.59 (1.30 – 1.96) | <0.001 | 1.50 (1.22 – 1.84) | **<0.001** |
| Female sex |  | 1 | - | 1 | - |
| Male sex | 104/1293 (8.04) | 1.17 (0.93 – 1.47) | 0.18 | 1.10 (0.87 – 1.39) | 0.43 |
| Age |  | - | <0.001* | - | <0.001* |
| Age 16-24 | 47/456 (10.3) | 1 | - | 1 | - |
| Age 25-34 | 152/1878 (8.1) | 0.77 (0.54 – 1.08) | 0.13 | 0.72 (0.51 – 1.02) | 0.069 |
| Age 35-44 | 102/1371 (7.4) | 0.70 (0.49 – 1.0) | 0.054 | 0.69 (0.48 – 0.99) | **0.044** |
| Age 45-54 | 71/1169 (6.1) | 0.56 (0.38 – 0.83) | 0.003 | 0.57 (0.38 – 0.83) | **0.004** |
| Age 55-64 | 32/721 (4.4) | 0.40 (0.25 – 0.64) | <0.001 | 0.44 (0.27 – 0.70) | **0.001** |
| Age 65-74 | 6/95 (6.3) | 0.59 (0.24 – 1.4) | 0.24 | 0.66 (0.27 – 1.59) | 0.34 |
| Age 75+ | 0/7 (0) | - | - | - | - |
| Job role |  | - | 0.0042* | - | 0.098* |
| Administrative | 19/412 (4.61) | 1 | - | 1 | - |
| Nursing staff | 261 / 3471 (7.52) | 1.68 (1.04 – 2.71) | 0.033 | 1.52 (0.94 – 2.46) | 0.088 |
| Junior doctor | 10/118 (8.47) | 1.92 (0.87 – 4.24) | 0.11 | 1.43 (0.64 – 3.23) | 0.39 |
| Consultant | 10/174 (5.75) | 1.26 (0.57 – 2.77) | 0.56 | 1.19 (0.53 – 2.68) | 0.42 |
| Healthcare assistant | 36/319 (11.29) | 2.63 (1.48 – 4.68) | 0.001 | 2.06 (1.14 – 3.71) | **0.016** |
| Theatre staff | 3/24 (12.5) | 2.95 (0.81 – 10.78) | 0.10 | 2.40 (0.65 – 8.87) | 0.19 |
| Physiotherapist | 9/84 (10.71) | 2.48 (1.08 – 5.69) | 0.032 | 1.82 (0.78 – 4.24) | 0.16 |
| Domestic and porter | 4/27 (14.81) | 3.60 (1.13 – 11.44) | 0.030 | 3.45 (1.07 – 11.2) | **0.039** |
| Other | 58/1068 (5.43) | 1.19 (0.67 – 2.02) | 0.53 | 1.06 (0.62 – 1.80) | 0.85 |
| Ethnicity | - | - | <0.001* | - | <0.001* |
| White | 275/4467 (6.16) | 1 | - | 1 | - |
| Black | 22/120 (18.33) | 3.42 (2.12 – 5.52) | <0.001 | 3.42 (2.12 – 5.53) | **<0.001** |
| Asian | 85/771 (11.02) | 1.89 (1.46 – 2.44) | <0.001 | 1.69 (1.30 – 2.19) | **<0.001** |
| Chinese | 5/78 (6.41) | 1.04 (0.42 – 2.60) | 0.93 | 1.04 (0.42 – 2.60) | 0.94 |
| Mixed | 2/68 (2.94) | 0.46 (0.11 – 1.90) | 0.28 | 0.42 (0.10 – 1.71) | 0.23 |
| Other | 9/97 (9.28) | 1.56 (0.78 – 3.12) | 0.21 | 1.36 (0.68 – 2.74) | 0.38 |
| Not stated | 12/96 (12.5) | 2.18 (1.17 – 4.04) | 0.013 | 2.10 (1.13 – 3.90) | **0.019** |

\*p value for the likelihood ratio test for the overall effect of variable

### Occupation

On univariate analysis, a number of HCW roles were associated with greater risk of the detection of SARS-CoV-2 antibodies. Nursing staff (OR 1·68 [95% CI 1·04 – 2·71]; *p*=0·033), healthcare assistants (HCAs) (OR 2·63 [95% CI 1·48 – 4·86]; *p*=0·001) physiotherapists (OR 2·48 [95% CI 1·08 – 5·69]; p=0·032) and porters and domestic staff (OR 3·60 [95% CI 1·13 – 11·44]; *p*=0·03) all displayed a higher risk compared to administrative staff (Table 2), who had the lowest seroprevalence at 4·6% (19/412). Security staff at CUH are employed by a third-party contractor and did not attend the staff serology testing clinic.

### Division/department

Staff departments at CUH are separated into six “Divisions” (Table 3). Risk differed significantly by division (*p* value for the likelihood ratio test of the overall effect of division <0·001). Staff working within Division C (primarily acute medicine, medical sub-specialities and Emergency Department) had a significantly higher seroprevalence (OR 2·8 [95% CI 1·82 – 4·33] *p*<0·0001) compared to staff working in Division E (paediatrics and maternity), who had the lowest seroprevalence (5·0%; 38/741). Staff working specifically in the ICUs had a seroprevalence of 6·33% (10/158), and staff working specifically in the Emergency Department had a seroprevalence of 9·1% (9/99). However, neither of these staff groups had significantly different seropositivity using univariate analysis (*p*>0·1 in both groups) compared to non-ICU and non-Emergency Department staff respectively.

**Table 3:**
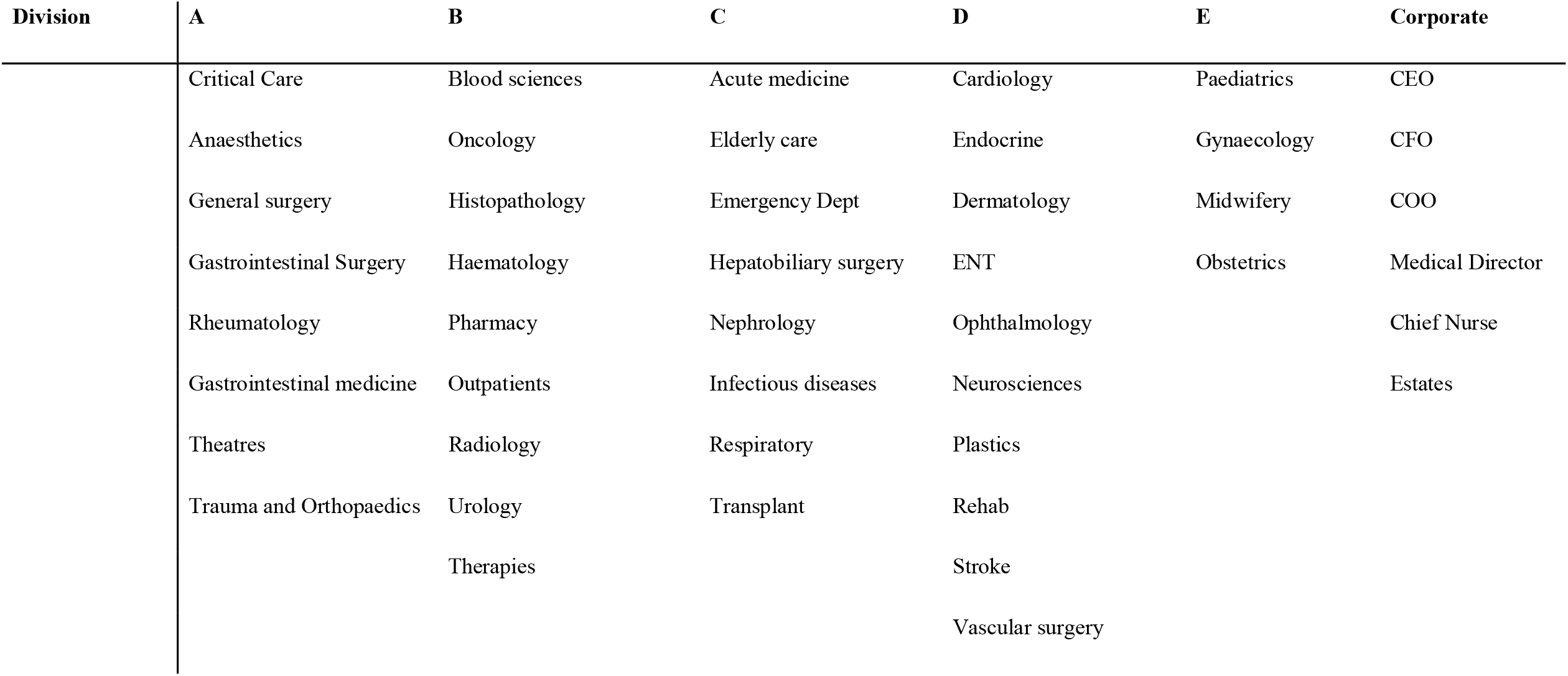
CUH departments by division

| Division | A | B | C | D | E | Corporate |
| --- | --- | --- | --- | --- | --- | --- |
|  | Critical Care | Blood sciences | Acute medicine | Cardiology | Paediatrics | CEO |
|  | Anaesthetics | Oncology | Elderly care | Endocrine | Gynaecology | CFO |
|  | General surgery | Histopathology | Emergency Dept | Dermatology | Midwifery | COO |
|  | Gastrointestinal Surgery | Haematology | Hepatobiliary surgery | ENT | Obstetrics | Medical Director |
|  | Rheumatology | Pharmacy | Nephrology | Ophthalmology |  | Chief Nurse |
|  | Gastrointestinal medicine | Outpatients | Infectious diseases | Neurosciences |  | Estates |
|  | Theatres | Radiology | Respiratory | Plastics |  |  |
|  | Trauma and Orthopaedics | Urology | Transplant | Rehab |  |  |
|  |  | Therapies |  | Stroke |  |  |
|  |  |  |  | Vascular surgery |  |  |

### Ethnicity

We observed substantial heterogeneity in the proportion of seropositivity between self-reported ethnic groups (Table 2). Staff identifying as White had an overall seropositivity rate of 6·1%. In comparison, Asian staff (Indian/Pakistani/Bangladeshi/Other Asian) and Black staff (Black African/Black Caribbean/Other black) had a seroprevalence of 11·0% (85/771) and 18·3% (22/120), respectively. White staff were more likely to have reported symptoms than Asian or Black staff (29%, 28% and 19% respectively). Despite this, seroconversion following symptoms consistent with COVID-19 was significantly higher in Black staff (*p*=0·002) and in Asian staff (*p*<0·001) compared to white staff; 41% (9/22), 26·6% (54/203), and 14·1% (148/1052) of staff had SARS-CoV-2 antibodies after reporting consistent symptoms in Black, Asian and White staff, respectively. The proportion of staff reporting having worked in a COVID-19 area was 38·5%, 60·3% and 32·1% for Black, Asian and White staff, respectively.

### Multivariable analyses

After describing several variables associated with SARS-CoV-2 seropositivity in a univariate analysis we included these variables to assess the risk associated with age, sex, ethnicity, job role and COVID-working status in a multivariable model. Increasing age remained protective for seropositivity on multivariable analysis (aOR 0·85 per 10 years increase in age [95% CI 0·78 – 0·93]; *p*<0·001). The aOR of having detectable SARS-CoV-2 antibodies in those that reported working in COVID-19 areas was 1·50 (95% CI 1·22 – 1·84; *p*<0·0001). Nursing staff and physiotherapists were no longer significantly associated with seropositivity on multivariable analysis (Table 2, Figure 3). HCAs remained at a significantly higher risk of being seropositive (aOR 2·06 [95% CI 1·14 – 3·71 – 2·4]; *p*=0·016), as were domestic and portering staff (aOR 3·45 [95% CI 1·07 – 11·2]; *p*=0·039). In a separate multivariable model including Division (Table 4), staff working in Division C (acute medicine, medical sub-specialities and Emergency Department) had an aOR of 2·07 (95% CI 1·31 – 3·25) for seropositivity (*p*=0·002) after controlling for age, sex, ethnicity, job role and COVID-19 area working.

**Table 4:** Odds ration (OR) and adjusted odds ratio (aOR) of seropositivity by Division.

| Variable | Seropositivity n (%) | Unadjusted OR (96% CI) | *p* value | Adjusted<sup>a</sup> OR (95% CI) | *p* value |
| --- | --- | --- | --- | --- | --- |
| Division |  | - | <0.001* | - | **0.0068\*** |
| A | 65/808 (8.0) | 1.62 (1.07 – 2.45) | **0.022** | 1.16 (0.74- 1.80) | 0.52 |
| B | 70/1211 (5.8) | 1.14 (0.76 – 1.70) | 0.54 | 1.08 (0.71 – 1.66) | 0.71 |
| C | 55/417 (13.2) | 2.81 (1.82 – 4.33) | **<0.001** | 2.07 (1.31 – 3.25) | **0.002** |
| D | 43/616 (7.0) | 1.39 (0.89 – 2.18) | 0.15 | 1.26 (0.80 – 1.99) | 0.33 |
| E | 38/741 (5.1) | 1 | - | 1 | - |
| Corporate | 26/445 (5.8) | 1.15 (0.69 – 1.91) | 0.60 | 1.24 (0.73 – 2.12) | 0.43 |

<sup>a</sup>Adjusted for age, sex, ethnicity, job role and working location
\**p* value for the likelihood ratio test for the overall effect of variable

**Figure 3:**
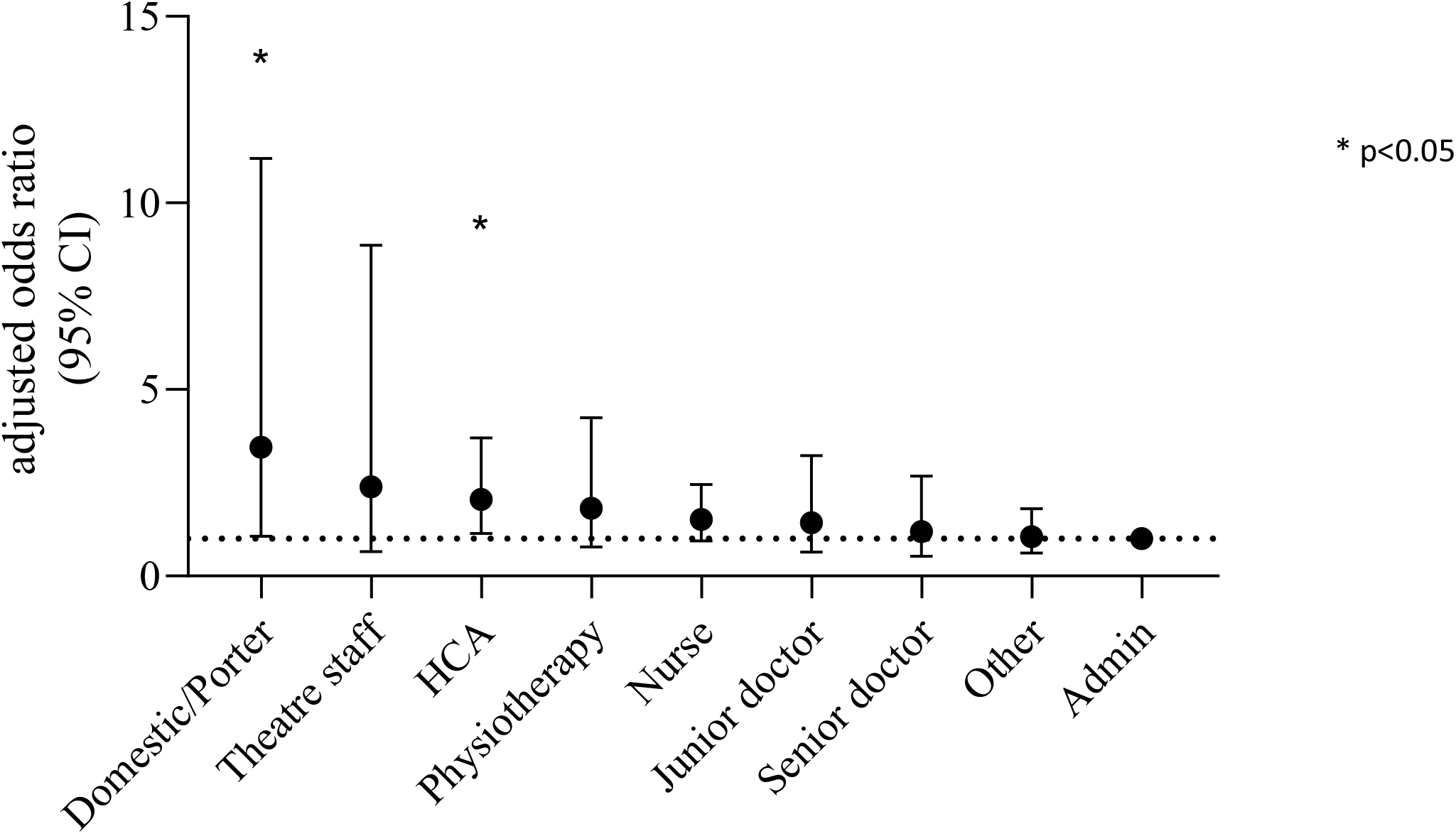
Adjusted odds ratio for SARS-CoV-2 seropositivity according to job role.

Ethnicity remained strongly associated with seropositivity (Table 2, Figure 4). The aORs in Asian and black staff in the multivariable model were 1·69 (95% CI 1·30 – 2·19; *p*<0·0001) and 3·42 (95% CI 2·12 – 5·53; *p*<0·0001), respectively (Table 2). There was no significant evidence that the effect of ethnicity was modified by COVID working location (*p* value of interaction 0·96), and we also observed a similar increase in risk associated with ethnicity when data were stratified by CUH COVID-19 working location. For Asian staff, the aOR for seroconversion was 1·59 (95% CI 1·09 – 2·32; *p*=0·016) for those working in COVID-19 areas compared to 1·76 (95% CI 1·21 – 2·55; *p*=0·003) for those not working in COVID-19 areas. For black staff the aOR for seroconversion when working in COVID-19 areas was 3·91 (95% CI 1·78 – 8·59; *p*=0·001), compared to 3·06 (95% CI 1·65 – 5·64; p<0·001) who reported working in a non-COVID-19 area.

**Figure 4:**
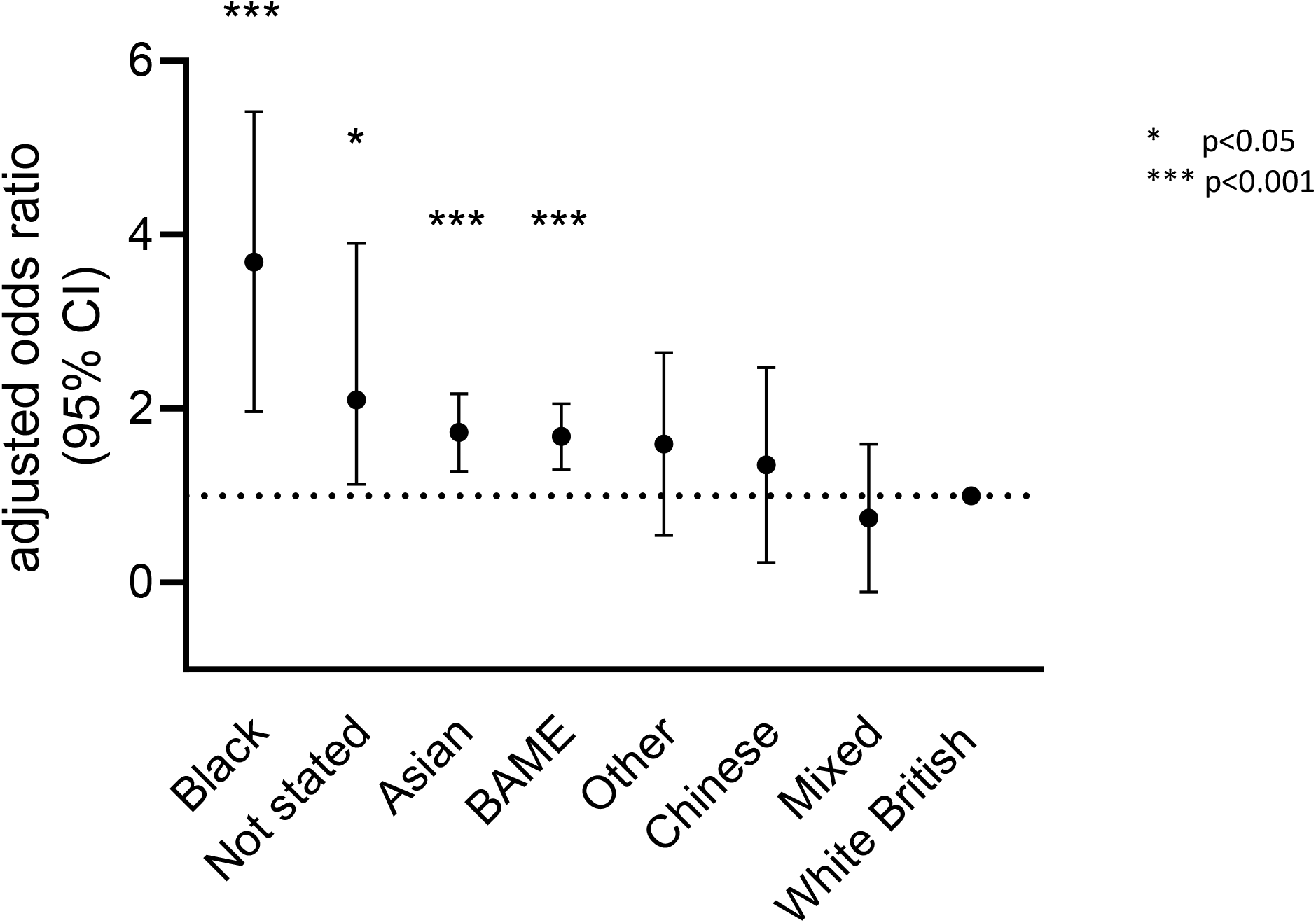
Adjusted odds ratio for SARS-CoV-2 seropositivity according to ethnic group.

The aOR for seropositivity in staff self-reporting as BAME (as a binary variable compared to white staff in a separate multivariable model) was 1·65 (95% CI 1·32 – 2·07; *p*<0·0001) after controlling for age, sex, job role and COVID-19 working location. For staff self-reporting as BAME, the aOR for seroconversion was 1·59 (95% CI 1·13 – 2·23; *p*=0·007) for those working in COVID-19 areas and 1·68 (95% CI 1·23 – 2·39; *p*=0·001) for those who reported not working in a COVID-19 area during the epidemic.

### Symptoms

Participants were asked about any symptoms consistent with COVID-19 since February 2020. Critically, seroprevalence was significantly higher in the group with symptoms (17·2%; 266/1,548) in comparison to those without symptoms (3·1%; 117/3827, *p*<0·0001). Almost 31% (126/410) of seropositive HCWs reported not having any symptoms consistent with COVID-19 since February 2020. After adjusting for age, sex and ethnicity, the aOR of seropositivity was 6·97 (95% CI 5·54 – 8·78; *p*<0·0001) in the group who reported prior symptoms compared to those who did not. The loss of the sense of taste or smell was the strongest predictor of seropositivity on univariate analysis; however, only 44% (154/351) of those reporting the loss of taste or smell were seropositive (Table 5). In a multivariable logistic regression model containing all collected symptoms (Table 5), loss of sense of taste or smell (aOR 7·85 [95% CI 5·79 – 10·65], *p*<0·0001), myalgia (aOR 1·71 [95% CI 1·18 – 2·48], *p*<0·0005) and fever (aOR 1·44 [95% CI 1·02 – 2·04], *p*<0·038) were the only symptoms positively associated with seropositivity. Notably, reporting a sore throat at the time of symptoms was negatively associated with seropositivity (aOR 0·7 [95% CI 0·50 – 0·99], p=0·043) in the multivariable model.

**Table 5:**
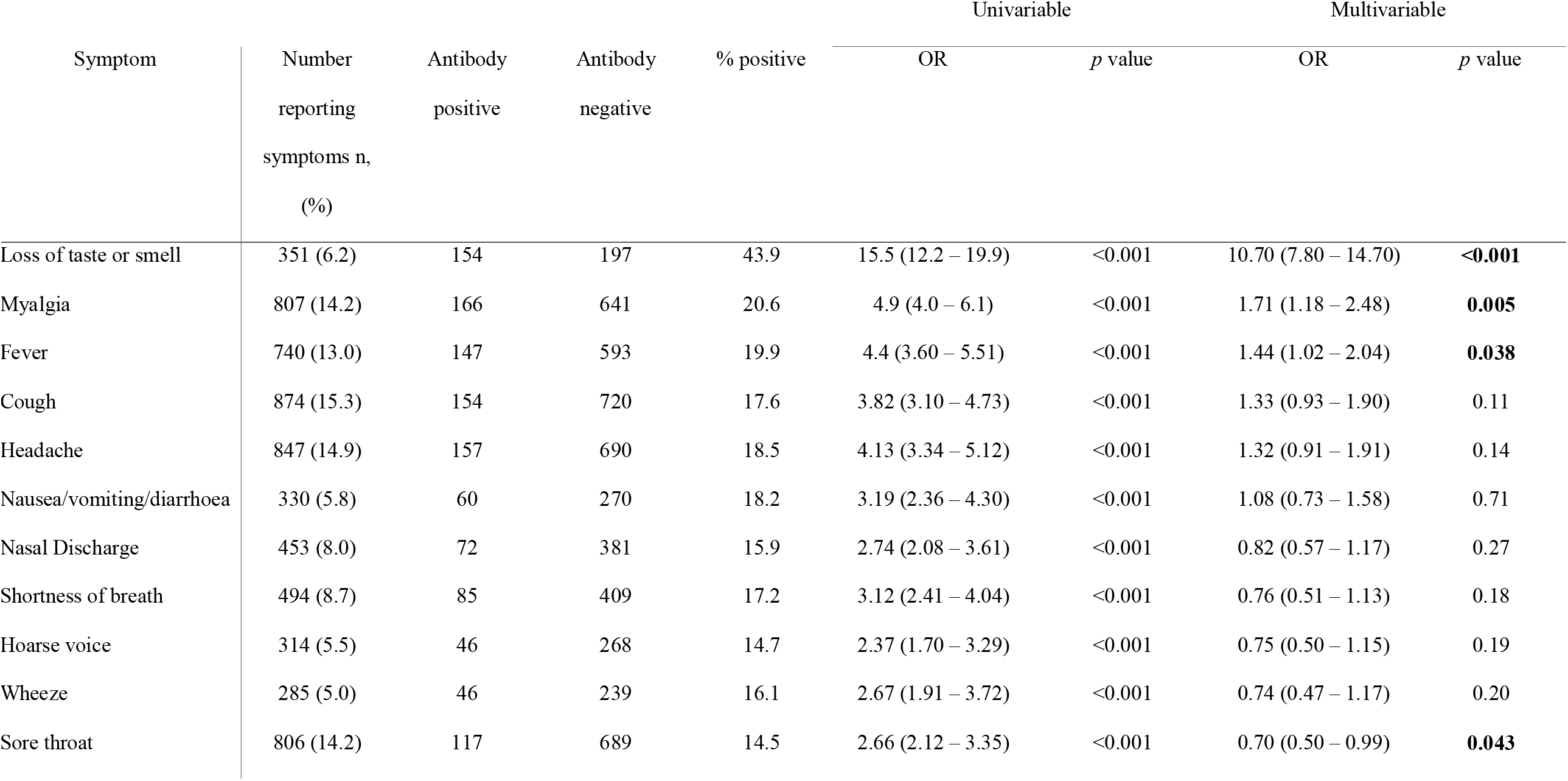
Unadjusted odds ratio (OR) and adjusted odds ratio (aOR) of SARS-CoV-2 seropositivity by reported symptoms

| Symptom | Number<br>reporting<br>symptoms n,<br>(%) | Antibody<br>positive | Antibody<br>negative | % positive | Univariable |  | Multivariable |  |
| --- | --- | --- | --- | --- | --- | --- | --- | --- |
|  |  |  |  |  | OR | *p* value | OR | *p* value |
| Loss of taste or smell | 351 (6.2) | 154 | 197 | 43.9 | 15.5 (12.2 – 19.9) | <0.001 | 10.70 (7.80 – 14.70) | **<0.001** |
| Myalgia | 807 (14.2) | 166 | 641 | 20.6 | 4.9 (4.0 – 6.1) | <0.001 | 1.71 (1.18 – 2.48) | **0.005** |
| Fever | 740 (13.0) | 147 | 593 | 19.9 | 4.4 (3.60 – 5.51) | <0.001 | 1.44 (1.02 – 2.04) | **0.038** |
| Cough | 874 (15.3) | 154 | 720 | 17.6 | 3.82 (3.10 – 4.73) | <0.001 | 1.33 (0.93 – 1.90) | 0.11 |
| Headache | 847 (14.9) | 157 | 690 | 18.5 | 4.13 (3.34 – 5.12) | <0.001 | 1.32 (0.91 – 1.91) | 0.14 |
| Nausea/vomiting/diarrhoea | 330 (5.8) | 60 | 270 | 18.2 | 3.19 (2.36 – 4.30) | <0.001 | 1.08 (0.73 – 1.58) | 0.71 |
| Nasal Discharge | 453 (8.0) | 72 | 381 | 15.9 | 2.74 (2.08 – 3.61) | <0.001 | 0.82 (0.57 – 1.17) | 0.27 |
| Shortness of breath | 494 (8.7) | 85 | 409 | 17.2 | 3.12 (2.41 – 4.04) | <0.001 | 0.76 (0.51 – 1.13) | 0.18 |
| Hoarse voice | 314 (5.5) | 46 | 268 | 14.7 | 2.37 (1.70 – 3.29) | <0.001 | 0.75 (0.50 – 1.15) | 0.19 |
| Wheeze | 285 (5.0) | 46 | 239 | 16.1 | 2.67 (1.91 – 3.72) | <0.001 | 0.74 (0.47 – 1.17) | 0.20 |
| Sore throat | 806 (14.2) | 117 | 689 | 14.5 | 2.66 (2.12 – 3.35) | <0.001 | 0.70 (0.50 – 0.99) | **0.043** |

### Seroconversion after positive SARS-CoV-2 PCR

From 5,991 enrolled participants, 2,825 (47%) reported having had a SARS-CoV-2 PCR test between February 2020 and the time of blood sampling, primarily through the CUH HCW testing programme. Of these, 51 (2·05%) tested PCR positive for SARS-CoV-2 RNA, 47 had detectable SARS-CoV-2 antibodies, and four had no detectable SARS-CoV-2 antibodies. The median SARS-CoV-2 PCR Ct value in those who seroconverted was 30 (IQR 24 – 34), in comparison to 36 (IQR 35·5 – 37) in those who did not seroconvert (*p*=0·006). The four staff who had previously tested SARS-CoV-2 PCR positive and were antibody negative all reported having symptoms consistent with COVID-19 infection at the time of PCR testing, although none reported the loss of taste or smell. Nine (18%) of the staff previously testing SARS-CoV-2 PCR positive, and who were antibody positive, were asymptomatic at the time of PCR testing.

## Interpretation

In this comprehensive assessment of factors associated with seropositivity for SARS-CoV-2 antibodies in HCWs at a large UK tertiary referral centre we were able to identify key at-risk occupational groups. Specifically, staff working in areas where patients with confirmed SARS-CoV-2 infection are cared for, those employed as HCA or domestic and portering staff, those of younger age, and those working in acute medicine or a medical sub-speciality were more likely to have SARS-CoV-2 antibodies. A reduced risk of SARS-CoV-2 seropositivity was associated with White ethnicity, being employed in an administrative role or working in paediatric or maternity services, and belonging to an older age group.

We found that the seroprevalence of SARS-CoV-2 antibodies in staff working in non-COVID facing areas was slightly higher (6·16%) than in the general population in the East of England (5·0%)^21^ and comparable to the national prevalence (6·0%)^21^. This is in keeping with previous retrospective serological HCW studies reporting relatively low seroprevalences in Germany (1·6%)^22^, Wuhan (3·8%)^23^ and Belgium (7·6%)^24^. Amongst Asian staff working at CUH the seroprevalence was also comparable to East of England data (10·5% vs 10·1%, respectively), as was the seroprevalence amongst Black staff at CUH compared to regional data (18% vs 15%, respectively)^21^. Overall, we observed significantly higher seroprevalence in all BAME staff compared to White staff, and to a greater extent in Black and Asian staff specifically. These differences have been observed nationally and are not unique to HCWs. The finding that the increased risk associated with BAME staff was not influenced by COVID-19 area working, as well as the ethnic differences in symptomatic seroconversion rates, demonstrates that the increased prevalence of antibodies in BAME HCWs cannot be accounted for by purely occupational factors.

In staff who were previously SARS-CoV-2 PCR positive, 92% (47/51) had detectable antibodies. There was a significant difference in Ct values between those who did and did not seroconvert. A potential explanation for this difference is that higher viral loads may be required to generate a sustained antibody response. Alternatively, a false positive SARS-CoV-2 PCR result or the detection of viral nucleic acid without infectious virus would also explain a lack of seroconversion.

Consistent with previous studies, we demonstrate that whilst reporting prior symptoms consistent with COVID-19 increased the chances of seropositivity, differentiating previous COVID-19 infection from other common respiratory tract infections based on symptoms alone is unreliable^6^. Interestingly, the only symptoms that significantly predicted seropositivity on a multivariable logistic regression model were the loss of sense of taste or smell, myalgia and fever. Prior reporting of cough or shortness of breath were not good predictors of the presence of SARS-CoV-2 antibodies in a multivariable model. These data also reiterate previous findings that asymptomatic SARS-COV-2 infection amongst healthcare workers is common, with 31% of seropositive staff having never reported consistent symptoms, and 18% of PCR positive staff never having reported consistent symptoms. These data highlight the importance of the contribution of the asymptomatically infected population to the spread of the disease^25,26^. Consequently, asymptomatic screening of staff in healthcare settings should become a component of routine disease survelliance^6,18,27^.

We acknowledge several limitations to our study. Variables such as ethnicity, COVID-working location and job role were self-reported; however, we have no reason to think these variables were party to recall bias and it is unlikely to impact on the results to any large degree. The proportion of staff reporting being of Black ethnicity was relatively small, although the proportion of BAME staff is consistent with the wider NHS workforce, and our conclusions are therefore broadly generalisable. The terminology and designation of COVID-facing clinical areas was an evolving factor throughout the course of the epidemic and is likely to be variable between hospital trusts and regions, as will the re-distribution of workforces and workflows through hospitals. Additionally, there will be heterogeneity of infection rates and admission pressures between different regions and between different hospitals within the same regions that may influence HCW exposure to infection differently. Consequently, this variation between practices may impact the specific risk factors assessed in this study to varying extents between different healthcare trusts. We were also unable to assess the use of PPE and adherence to PPE protocols in this study design. The selected assay may have reduced sensitivity in individuals who generated robust antibody responses to other SARS-COV-2 antigens or those producing low affinity antibodies during early disease. Similar considerations apply to other commercial assays^28^, and a recent comparison demonstrated assay equivalence with the selected platform having higher accuracy^19^, and an independently reported sensitivity and specificity of 98·1% (95% CI 96·6 – 99·1) and 99·1% (95% CI 99·4 – 100) respectively^19^. We also note that symptom data were recorded retrospectively, and may have been subject to recall bias.

Our study provides new information on the risk factors for SARS-COV-2 infection and antibody response in healthcare facilities. We found that the occupational exposure to SARS-COV-2 is heterogenous across job roles, hospital department, and ethnicity. It is clear that HCWs on the frontline of the COVID-19 pandemic require more protection from occupational exposure with accurate stratification of risk factors to develop mitigation strategies. The association with ethnic group is concerning and a deeper understanding of the societal and/or genetic factors predisposing the BAME population to SARS-COV-2 infection and seroconversion is needed.

## Data Availability

Requests for de-identified data will be considered upon request.

## Acknowledgements

We thank NIHR BioResource volunteers for their participation, and gratefully acknowledge NIHR BioResource centres, NHS Trusts and staff for their contribution. We thank the National Institute for Health Research, NHS Blood and Transplant, and Health Data Research UK as part of the Digital Innovation Hub Programme. The views expressed are those of the author(s) and not necessarily those of the NHS, the NIHR or the Department of Health and Social Care. The authors also thank the Occupational Health Department at CUH for facilitating sample and data collection during the staff serology testing clinic.

